# Multiscale Spatial Characterization of Glioma MRI with MUSE and Radiomics

**DOI:** 10.64898/2026.10.27.26366624

**Authors:** Marianna Miola, Simone Cammarasana

## Abstract

**Purpose:** Characterizing glioma heterogeneity requires measurements that retain the physical scale and orientation of intratumoral intensity dependence. We evaluate the original 3D variographic field produced by the `MUSE` tool (Modeling Uncertainty as a Support for the Environment) alongside directional radiomics and independent image-based molecular classification.

**Methods:** We analyzed postcontrast T1-weighted magnetic resonance images and enhancing-tumor segmentations from the public UCSF-PDGM collection. Verified `MUSE` outputs were available for 416 independent baseline patients, of whom 375 had paired directional gray-level co-occurrence matrix (GLCM) measurements. Each `MUSE` field retained 328 original experimental directional variograms with physical lag distances, semivariances, orientations, and voxel-pair support. Agreement was assessed at aligned offsets and through between-tumor comparisons. Patient-level permutation tests evaluated associations with isocitrate dehydrogenase (IDH) status on 2,508 native direction-lag coordinates. A standalone 3D residual convolutional neural network (CNN) provided a separate IDH classification benchmark.

**Results:** At 9,371 geometrically aligned offsets, native `MUSE` semivariance and half GLCM contrast correlated strongly (Pearson *r* = 0.954). Across 31,599 eligible between-tumor comparisons, representation-level dissimilarities showed Spearman *ρ*_S_ = 0.862, with sensitivity to GLCM feature family and long-offset sampling support. In 364 grade-4 tumors, including 24 IDH-mutant cases, the original `MUSE` field was associated with IDH after conventional adjustment (global permutation *p* = 0.00050), but the association was attenuated after primary directional GLCM adjustment (*p* = 0.256). The independent CNN achieved an out-of-fold IDH ROC-AUC of 0.792 and PR-AUC of 0.414 across 416 patients. Native `MUSE` examples revealed scale- and direction-dependent spatial behavior, including distances beyond those sampled by the GLCM protocol.

**Conclusion:** Variograms by `MUSE` provide a physically indexed description of tumor spatial heterogeneity that can accompany radiomic analysis and CNN classification. The observed statistical overlap is substantial, and incremental clinical discrimination beyond directional GLCM features remains unestablished.

## 1. Introduction

Diffuse gliomas exhibit spatially heterogeneous enhancement, cellularity, vascularity, and treatment-relevant molecular characteristics. Magnetic resonance imaging (MRI) provides a noninvasive representation of several aspects of tumor phenotype; the enhancing-tumor (ET) compartment in postcontrast T1-weighted (T1c) imaging offers a spatially defined region for quantitative analysis. Molecular classification and integrated diagnosis are established components of contemporary glioma assessment [1]. Their relationship to imaging heterogeneity motivates methods that retain the spatial organization of tumor signal.

Radiomics extracts morphological, intensity, and texture measurements from segmented images and has become a widely used paradigm for quantitative imaging research [2, 6]. Gray-level co-occurrence matrices (GLCMs), in particular, encode the joint distribution of quantized intensities at specified spatial offsets [7]. Directional and multiscale GLCMs contain rich information, although standard feature pipelines often aggregate directional measurements for comparability and statistical learning. The resulting summaries can be informative for classification and prognosis while leaving the dependence on the underlying physical separation and orientation less directly visible in an individual patient.

Geostatistical analysis offers a physically indexed representation of dependence between sampled locations. An experimental semivariogram expresses the squared change in a spatially indexed field as a function of physical separation and direction [8]. The present biomedical study employes the open-source `MUSE` tool [9] for computing 3D experimental variography to segmented glioma MRI, and evaluates the native fields against directional radiomics, molecular annotations, and independent image-based classification. Applied to a segmented tumor, the experimental output can be examined as a family of semivariance curves over physically measured lag distances and spatial orientations. This output describes the spatial arrangement of MRI intensities conditional on the segmented enhancing-tumor domain. Biological interpretations of these measurements require independent evidence.

`MUSE` and GLCM are expected to share substantial information. Under identical voxel-pair support and without quantization differences, the GLCM contrast is related algebraically to twice the semivariance. The present comparison quantifies agreement between the native representations, identifies the effect of finite sampling support, and examines how their spatial coordinates support interpretation of individual cases. In particular, the original 3D variogram field makes the scale- and direction-dependent behavior explicit, whereas individual GLCM measures also encode aspects of the full co-occurrence distribution, including entropy and energy.

We developed and evaluated an integrated radiomic–geostatistical workflow on the publicly available University of California San Francisco Preoperative Diffuse Glioma MRI (UCSF-PDGM) dataset [17, 18]. The technical archive comprised 416 unique baseline cases with verified `MUSE` results, and 375 were available for paired multiscale directional GLCM analysis. Across 31,599 eligible intertumor comparisons, the two representation-level dissimilarities showed a Spearman rank correlation (*ρ*_S_ = 0.862). At geometrically aligned offsets, `MUSE` semivariance and half GLCM contrast also agreed strongly (*r* = 0.954). Offset scale, voxel-pair support, and GLCM feature family influenced the measured degree of agreement. A patient-level, multiplicity-controlled analysis of the original direction–lag measurements found a clinical association with isocitrate dehydrogenase (IDH) status under conventional adjustment, but not after the primary directional GLCM adjustment.

An independent 3D convolutional neural network (CNN) extends this assessment to image-based deep learning. The network predicts IDH status from a tumor-centered MRI volume and its segmentation, while original `MUSE` measurements remain available for post-prediction spatial characterization. The study thereby reports image-based discrimination and physical description as two observational outcomes for the same tumors. The independent CNN achieved an out-of-fold receiver operating characteristic area under the curve (ROC-AUC) of 0.792 among 416 baseline cases, including 43 IDH-mutant tumors. On the 375 subjects shared with the directional radiomic analysis, its already generated predictions yielded ROC-AUC 0.808. The neural predictions and original `MUSE` spatial observations are reported as two separate outcomes for each selected tumor.

The contributions of this study concern three connected aspects of biomedical spatial characterization. First, we preserve and analyze the complete native experimental 3D `MUSE` field, including physical lag, direction, semivariance, and pair count, using the original `MUSE` files as the authoritative record. Second, we establish a geometry-aware comparison with oriented multiscale GLCMs and quantify the influence of sampling support on apparent representational differences. Third, we relate the spatial characterization to molecular annotations through cohort-level testing and predefined cases and report independent CNN classification as a benchmark for post-prediction spatial characterization. The study quantifies representational agreement and exploratory clinical association within a public imaging cohort.

## 2. Related Work

### 2.1 Quantitative MRI and medical-image radiomics

Texture analysis of images began with co-occurrence features describing intensity transitions at defined spatial offsets [7]. High-dimensional radiomics broadened quantitative tumor descriptions to include morphological, intensity, and filtered texture properties [2, 3]. Reproducible extraction software made large families of image biomarkers accessible across clinical datasets [6]. Subsequent reviews identified image normalization, feature definitions, validation design, and repeatability as determining factors in reproducible radiomic inference [4, 5]. The Image Biomarker Standardization Initiative established consensus feature definitions and quantitative benchmarks [19]. Imaging perturbation experiments evaluated the sensitivity of radiomics to segmentation and acquisition changes [20].

Neuro-oncology datasets provide a basis for evaluating these measurements under standardized clinical annotations. Expert-segmented glioma image collections enabled radiomic analysis of anatomical subregions [21]. Multimodal segmentation benchmarks supported systematic comparisons of brain-tumor processing algorithms [22]. Self-configuring neural segmentation provided an adaptable computational framework for diverse biomedical imaging tasks [23]. The UCSF-PDGM collection offers a complementary public resource with preoperative imaging, tumor segmentation, molecular characterization, and follow-up metadata [17, 18]. Established imaging-biomarker qualification frameworks distinguish measurable image characteristics from clinically validated biomarkers [24]. Bias-field correction and medical-image software platforms constitute complementary preprocessing and research infrastructure developments [31, 32].

#### Image-based deep learning for molecular classification

Deep residual architectures provide a standard reference for learning nonlinear representations from imaging data [36]. Preoperative MRI-based residual classification has been evaluated for isocitrate dehydrogenase genotype in low- and high-grade gliomas [37]. Fully automated deep-learning methods have also been developed for MRI-based IDH classification using tumor-related image information [38]. Segmentation-focused architectures, including self-configuring neural networks, provide upstream image-processing tools for defining anatomical regions of interest [23]. These methods evaluate learned predictive representations; explicit physical-direction and lag-dependent inspection is provided separately by the experimental variographic measurements in the present workflow.

### 2.2 Spatial modeling, directional dependence, and geometric signals

The experimental variogram characterizes separation-dependent variability in a regionalized field and has established theoretical foundations in spatial statistics [8]. Its direct use in a segmented MRI compartment preserves observed distance, direction, and voxel-pair support. `MUSE` provides a computational setting for evaluating such 3D spatial representations [9]. Earlier heterogeneous volumetric modeling combined geometric representation and spatial estimation for incomplete subsurface observations [10]. Spatial geochemical modeling formalized validation and uncertainty-aware inference for georeferenced fields [11]. Reproducible scientific computation was further addressed through traceable geoscientific workflow persistence [12].

Related work in geometric signal analysis provides a broader context for interpreting spatially organized measurements. Kernel-based sampling addressed signals over irregular geometric domains [13]. Localized spectral constructions analyzed shape-aware functions and multiscale geometric structure [14]. In biomedical signal processing, ultrasound denoising studies evaluated the preservation of anatomical information under learned image transformations [15]. Ultrasound super-resolution addressed reconstruction of spatial image detail and video sequences [16]. These developments concern related problems in spatial signals and geometry; the present study evaluates original geostatistical dependence measurements in MRI tumor masks.

### 2.3 Statistical reliability and clinical association

Statistical validation of imaging biomarkers requires attention to the observational unit, multiplicity, and model-selection uncertainty. Regression-based clinical survival analysis originates from proportional-hazards methods and their validation methodology [33, 34]. Multiple-testing adjustments control expected false discoveries across correlated exploratory findings [35]. Resampling and permutation procedures support inference while accounting for analysis design and joint test dependence [25, 26, 28]. Nested validation addresses bias introduced when model tuning and evaluation share observations [27]. Cross-scanner neuroimaging harmonization and related empirical-Bayes adjustments highlight additional potential sources of nonbiological imaging variation [29, 30]. The experiments reported here consequently preserve patient identity in uncertainty calculations and treat directional sampling support as an explicit component of the comparison protocol.

## 3. Materials and Methods

The analysis protocol links the public imaging collection, native three-dimensional experimental variograms, and oriented co-occurrence statistics through physical geometry and recorded spatial support. Cohort construction and measurement definitions are followed by the paired comparison, sampling-sensitivity, and molecular-association procedures.

### 3.1 Study cohort and imaging domain

The public UCSF-PDGM collection includes 501 imaging examinations corresponding to 495 unique patients, with 3D preoperative MRI, segmentations, molecular data, and follow-up metadata [17, 18]. We analyzed T1c intensities within the supplied enhancing-tumor mask. Seventy-five examinations had no ET voxels and were not applicable to ET-restricted variography. Of 426 ET-positive examinations, four did not complete the `MUSE` pipeline. The remaining 422 successful examinations included six follow-up acquisitions; retaining the baseline examination for each patient yielded 416 independent cases. A common dataset of 375 cases possessed the aligned normal-score radiomics and directional GLCM outputs required for the paired study (Fig. 2 and Table 1). Figure 1 displays the original postcontrast T1-weighted anatomical sections and corresponding ET segmentation of patient UCSF-PDGM-0531, selected for the directional case-level analysis.

**Table 1.** Analysis populations and original measurement inventories. Cohort counts refer to independent baseline subjects after the stated selection stages.

| Population or measurement | Count |
| --- | --- |
| Original UCSF-PDGM examinations | 501 |
| ET-absent examinations | 75 |
| ET-positive examinations | 426 |
| MUSE technical failures among ET-positive cases | 4 |
| Technically valid MUSE examinations | 422 |
| Independent MUSE baseline patients | **416** |
| Full-cohort native MUSE directional curves ($416 \times 328$) | 136,448 |
| Full-cohort native directional lag samples | 1,151,687 |
| Paired MUSE –GLCM baseline patients | **375** |
| Paired-cohort MUSE directional curves | 123,000 |
| Paired-cohort original directional lag samples | 1,075,217 |
| Grade-4 patients in native IDH localization | 364 |

**Figure 1:**
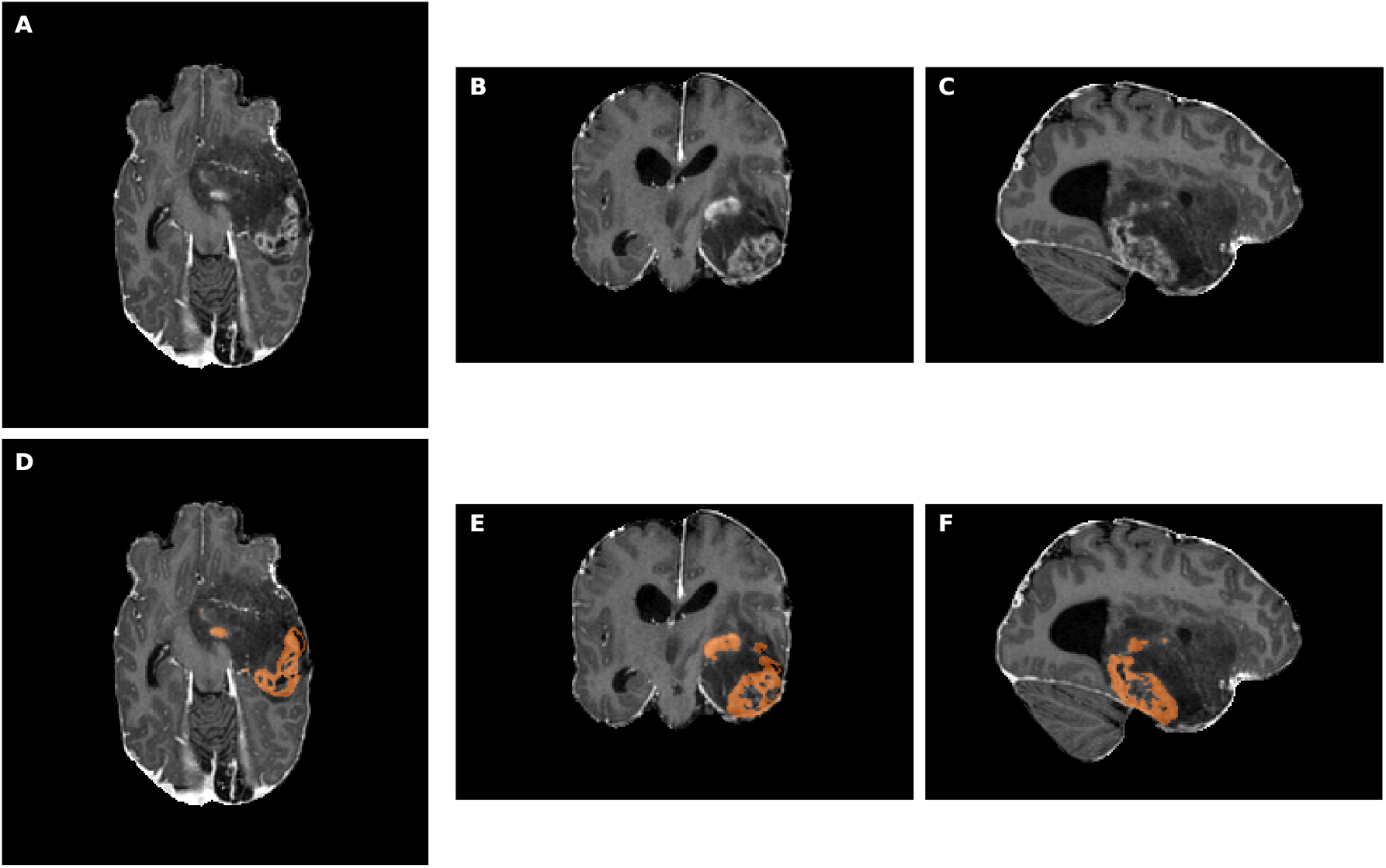
Postcontrast T1-weighted magnetic resonance images of patient UCSF-PDGM-0531. **(A–C)** Axial, coronal, and sagittal sections, respectively. **(D–F)** Corresponding sections with the enhancing-tumor (ET) segmentation superimposed in orange. Anatomical sections and ET masks are obtained from the original patient image and segmentation volumes.

**Figure 2:**
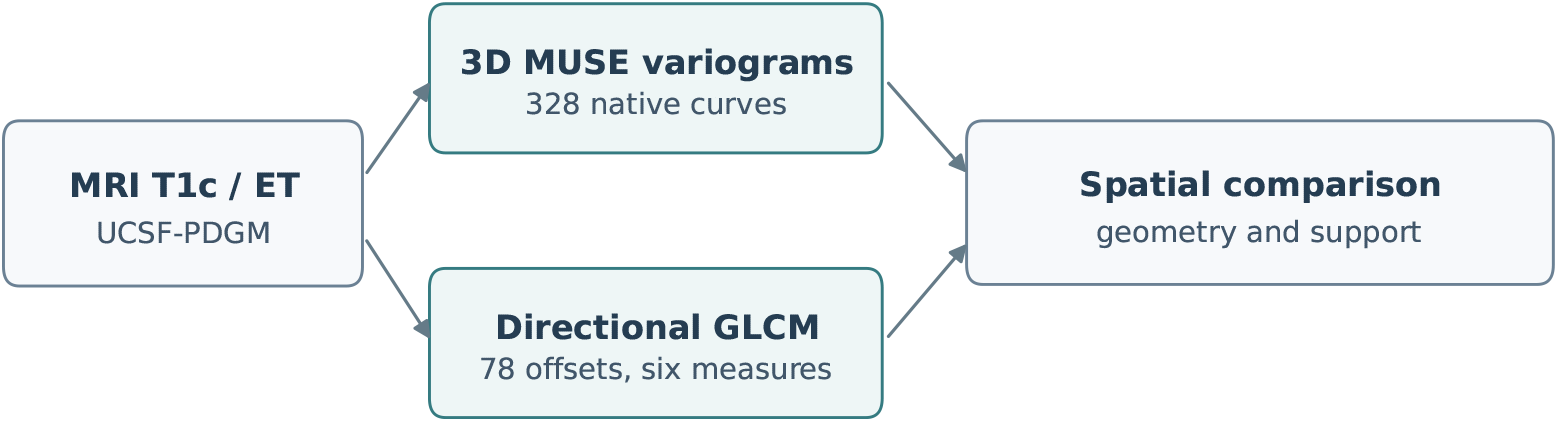
Workflow of the image-based analyses. Tumor-centered postcontrast T1-weighted MRI and enhancing-tumor segmentation define the common input domain (left). `MUSE` 3D experimental variograms and directional multiscale GLCM features are compared at supported physical separations (central and upper-right boxes). The line below the workflow identifies the independently evaluated residual CNN for IDH classification and the subsequent inspection of the `MUSE` field. Cohort inclusion counts appear in Table 1.

The original NIfTI geometries were canonicalized to RAS+ for correspondence with discrete GLCM offsets; canonicalization did not imply spatial resampling. Voxel dimensions in the paired cohort were isotropic at 1 mm and had no relevant obliquity in the audit. Intensities were transformed within each ET region using an ordinal normal-score transformation to reduce marginal intensity variation. In the GLCM implementation, ordinal ranks were transformed by a Gaussian quantile mapping and quantized with bin width 0.2. The native `MUSE` computation used the corresponding continuous normal-score intensity field; small differences between transformations and discrete versus continuous estimators remain possible.

### 3.2 Original MUSE and directional co-occurrence measurements

`MUSE` was executed using original normal-score ET voxel data with 3D directional variography (automatic plane generation, plane-step setting 20); the archived run sampled 82 planes and four directions per plane, producing 328 experimental directional curves per case. All downstream analyses used the previously archived `MUSE` computations. For sampled intensity *Z*(**x**) and a direction–lag pair set *P*_*θ,h*_, the experimental semivariance is

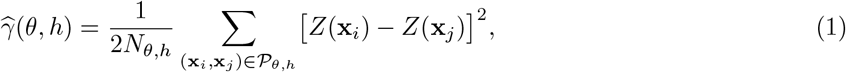

where *θ* identifies a 3D axial orientation, *h* is physical separation, *N*_*θ,h*_ is the number of contributing voxel pairs, and the pair set follows `MUSE` ‘s original angular and lag tolerances. Opposite unit vectors represent the same semivariogram axis. Equation 1 defines the experimental estimator used to interpret the original measurements. The primary data consist of the original lag *h*, semivariance 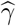, support *N*, plane identity and physical direction vector for every recorded point.

Direction vectors were reconstructed from the documented native plane azimuth, dip, and in-plane angle using the `MUSE` implementation, and verified against stored plane normals. Source JSON files were associated with SHA256 hashes; export and audit tables confirm their provenance. All retained values are original experimental measurements. The primary representation comprises the originally archived curves with their measured coordinates and support; fitted ellipsoids and scalar comparison indices are auxiliary outputs. Omnidirectional curves and geometric fits were retained as supporting representations. Figures 3 and 4 reproduce six original outputs from a T1c `MUSE` directional-analysis example supplied by the authors. The four plane-0 variograms contain their original experimental observations and fitted curves; the range diagram and the 3D ellipsoid are fitted geometric summaries of directional range estimates. The example is used to illustrate the interpretation of the software outputs. Its source images do not carry a cohort subject identifier, and it is therefore kept separate from the patient-specific results and inference in Section 4. The original sampled-direction display used in the patient comparisons is shown separately in Fig. 6. A between-patient distance or scale-band statistic is explicitly considered a *secondary comparison diagnostic*, not a new `MUSE` representation.

**Figure 3:**
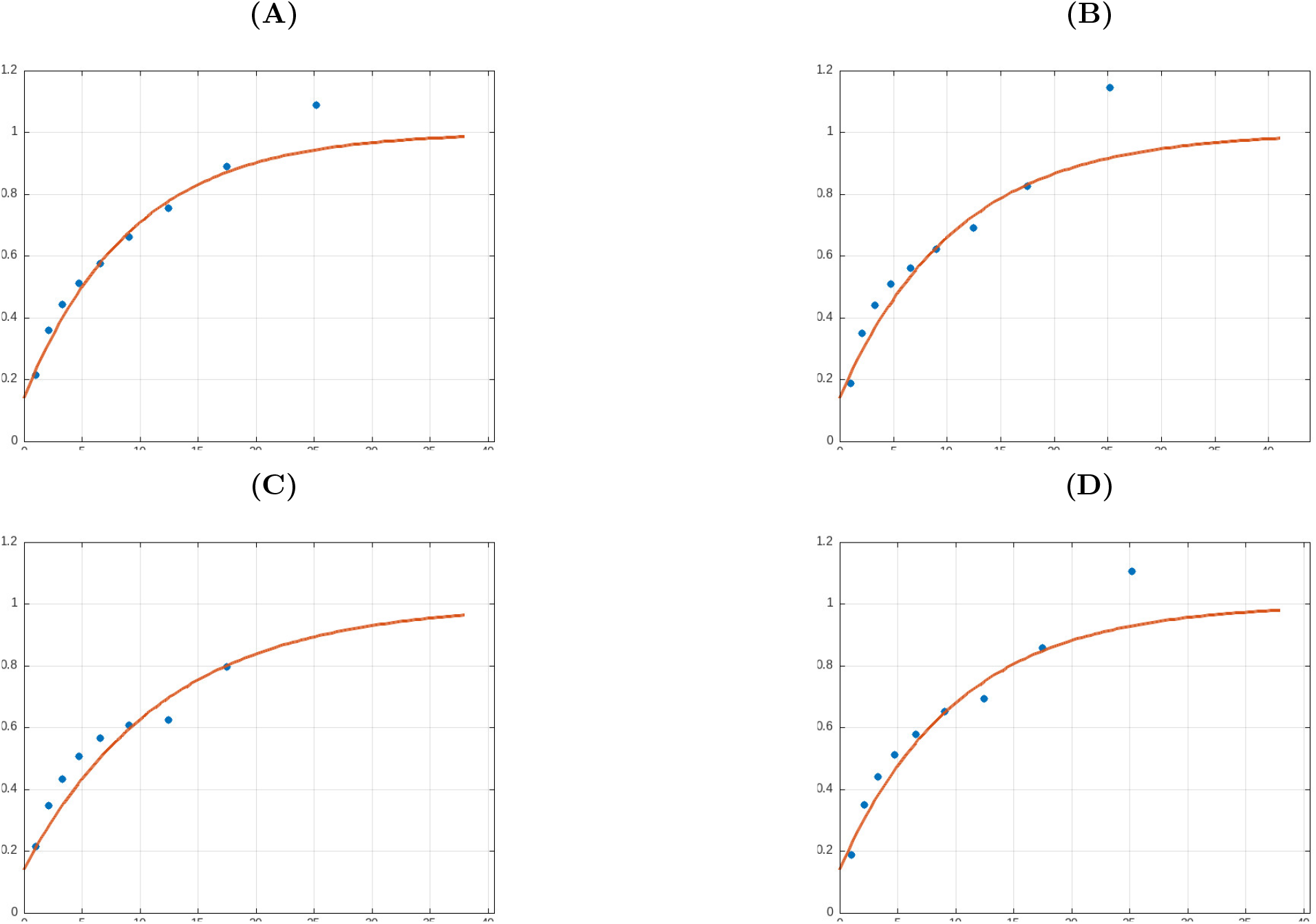
Original `MUSE` directional variograms in the four sampled directions of plane 0. Panels (A)–(D) correspond to direction indices 0, 1, 2, and 3, respectively. Each panel plots experimental semivariance (vertical coordinate) against the sampled lag distance (horizontal coordinate). Blue points are experimental observations and orange lines are fitted variogram curves.

**Figure 4:**
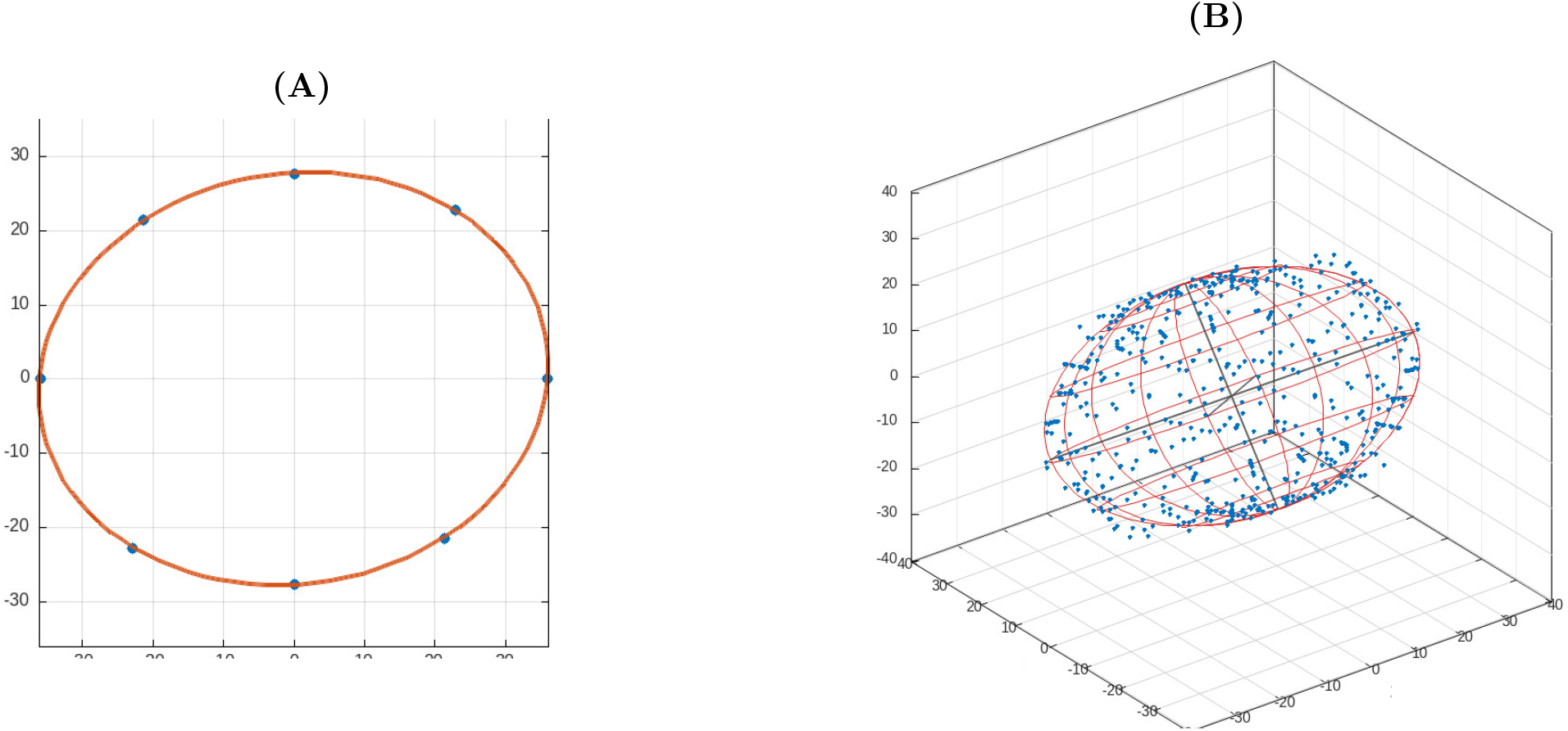
Geometric outputs of the `MUSE` directional analysis. (A) Fitted range diagram in plane 0, plotted in the local east (horizontal coordinate) and north (vertical coordinate) directions. Blue points mark estimated directional ranges and the orange contour shows their fitted planar ellipse. (B) 3D fitted anisotropy ellipsoid shown in its original east–north–vertical coordinate frame. The source output depicts directional points in blue, the ellipsoid mesh in red, and principal-axis segments in dark lines. The displayed fitted semi-axis lengths are 34.7, 31.5, and 23.0 in the original renderer’s coordinate units.

The four experimental semivariograms in Fig. 3 illustrate the lag-dependent structure recorded by `MUSE` for four directions in plane 0. The measured semivariance rises over short and intermediate separations and approaches a similar upper level across the displayed directions. The fitted curves facilitate an inspection of scale-dependent continuity, while the original points retain the variation caused by finite sampling support. The corresponding planar range diagram and 3D ellipsoid in Fig. 4 provide a geometric summary of fitted directional ranges. In the supplied illustration the reported 3D semi-axis lengths are 34.7, 31.5, and 23.0 in the renderer’s native coordinate units. The first two lengths are comparatively close, whereas the third is shorter, illustrating how a 3D fitted summary can reveal a different directional extent from a single planar section. These fits are used for visual interpretation; the quantitative experiments in this study continue to operate on the full original experimental direction–lag samples.

#### Directional multiscale GLCM and radiomic features

The complementary texture experiment considered 13 axial lattice-offset representatives at integer voxel steps *d* ∈ {1, 2, 3, 5, 8, 11}. At 1 mm isotropic spacing, the physical offsets have lengths 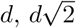, or 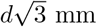, with a maximum of approximately 19.05 mm. The 78 orientation–step combinations yielded six GLCM measures each: contrast, dissimilarity, homogeneity, correlation, Shannon entropy, and energy (468 native oriented entries per patient). Offset-specific voxel-pair counts were archived; offsets supported by fewer than 100 pairs were excluded by the original extraction gate. We also retained orientation-invariant within-physical-length summaries and a previously computed set of 203 harmonized conventional and multiscale radiomic features [6]. All comparisons used the archived GLCM measurements and `MUSE` experimental output as fixed inputs.

For a normalized co-occurrence matrix *P*_***δ***_(*i, j*) at spatial offset ***δ***, GLCM contrast can be written as

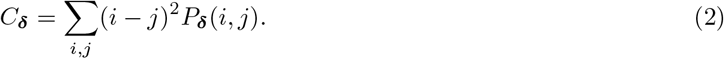

Here *i* and *j* are discrete gray-level indices, *P*_***δ***_(*i, j*) is their normalized joint frequency for voxel displacement ***δ***, and *C*_***δ***_ is contrast. When the co-occurrence entries represent the same continuous intensities, voxel pairs, and direction as the semivariogram estimator, *C*_***δ***_*/*2 coincides with the corresponding sample semivariance. With discrete gray levels, exact lattice offsets, symmetric counting, and different angular supports, this identity becomes an interpretive relationship and not a guaranteed numerical equality for the present two outputs. Equation 2 characterizes the second moment of the measured transitions. Entropy and energy also depend on the full co-occurrence distribution and are not determined by a second-order contrast alone.

### 3.3 Geometric alignment and representation comparison

A `MUSE` direction and a GLCM offset were compared using the axial angular distance arccos |**u**_MUSE_ *·* **u**_GLCM_| and the absolute difference in physical lag. The primary diagnostic allowed up to 7.5^°^ axial mismatch and a lag discrepancy bounded by max(0.35 mm, 0.05*h*), with at least 100 contributing native `MUSE` voxel pairs. A stricter diagnostic used 3^°^ and 0.1 mm. These tolerances identify geometrically comparable observations; they do not equate the estimators’ directional bandwidths. Cohort-level matching used all supported GLCM orientations and offsets, while recording the fraction without a geometrically qualified `MUSE` counterpart.

#### Between-tumor representation agreement

The representation-level comparison examined all original `MUSE` curves for pairs of patients by exact direction identity and by directly paired native lag samples within 0.1 mm, without interpolation. Pair eligibility additionally required a tumor-volume ratio no greater than 2, at least 65% available oriented GLCM entries, at least 400 matched original `MUSE` points over 160 or more directions, and at least 30% native `MUSE` observation coverage. A root-mean-square difference of the original matched semivariances served as an *index for comparing patients*; the full curves remained available for inspection. Directional GLCM distances were constructed from all 468 entries after feature-wise robust median/IQR scaling and evaluated alongside distances computed for each of the six GLCM families. Rank concordance was assessed using Spearman’s correlation across eligible patient pairs. The 31,599 overlapping patient pairs are statistically dependent, so no ordinary independent-pair significance test was used; leave-one-patient-out and patient-subset checks evaluated stability.

A second, pointwise analysis used the geometrically aligned `MUSE` and GLCM contrast measurements. Correlations were computed over aligned observations, within patients, and after removing patient-specific effects at identical physical GLCM offset length. Patient-cluster resampling was used for descriptive uncertainty. Because matched observations arise from different support sets, their numerical differences were not labeled measurement errors.

#### Support and sensitivity diagnostics

The number of independent spatial pairs decreases as tumor masks become smaller, irregular, or sparse at long offsets. We therefore repeated the GLCM distance comparison by masking already extracted oriented feature entries with fewer than 200, 300, 500, and 1,000 saved contributing pairs. `MUSE` gamma measurements were not changed. We also decomposed the squared robustly standardized distance by GLCM family and voxel step, tested representation subsets with and without entropy and energy, and checked whether the original candidate discordant tumor pairs persisted. These are sensitivity analyses of the selected representation, not alternative estimates of the true MRI texture.

### 3.4 Molecular association and illustrative case design

A separate clinical experiment evaluated isocitrate dehydrogenase (IDH) status within grade-4 tumors to restrict gross differences due to grade composition. The intersection of original `MUSE`, oriented GLCM, and clinical records included 364 grade-4 tumors (24 IDH-mutant, 340 IDH-wildtype). The original `MUSE` export contributed 1,046,436 examined lag observations. Using a minimum of 500 original voxel pairs per point and label-blind coverage of at least 260 patients, 2,508 original curve-index–physical-lag cells were eligible for inference. The analysis operated on experimental semivariance at those exact original coordinates and did not construct patient-level `MUSE` biomarkers or a surrogate field.

At each tested cell, semivariance was residualized against age, log ET volume, shape anisotropy, raw T1c mean and standard deviation, and sex. The primary additional texture adjustment added five principal components of the 468 original oriented GLCM features, with ten components examined as sensitivity. Principal component analysis was applied to the *GLCM control data only*; native `MUSE` was neither decomposed nor replaced by a fitted field. Cells entered testing according to label-blind coverage; patient-level labels were permuted 2,000 times within strata defined by age, tumor volume, and sex. Standardized signed cell scores contributed to a global mean-squared-score statistic. The maximum absolute permuted cell score provided family-wise error rate (FWER) adjustment over all 2,508 tested cells. This is an exploratory conditional permutation design; exchangeability within the strata and representativeness of the partial GLCM adjustment are assumptions, not guaranteed properties.

A previously completed secondary comparison of compact fitted `MUSE` summaries and radiomics was retained solely for context, including internally validated IDH and survival models. It is reported separately from the original-field experiment. It cannot establish or refute incremental clinical utility of the complete 328-curve field.

#### Outcome-independent illustrative case selection

A set of 15 opposite-IDH grade-4 case pairs was selected using radiomic proximity and conventional demographic/morphological constraints without screening native `MUSE` curves for favorable differences. The original export contained 83,875 semivariance observations spanning 9,512 complete experimental curves from 29 unique tumors, together with 468 oriented GLCM measures per tumor and their clinical metadata. The 15 pairs served as illustrative and negative-control candidates; matched IDH labels by construction cannot establish diagnostic association. Selected examples were displayed with the full original curves and original GLCM-offset values. When scale-band differences were tabulated, these were explicitly secondary descriptions of the displayed native measurements.

### 3.5 Independent convolutional classification benchmark

An independent 3D residual convolutional neural network was evaluated for patient-level classification of IDH-mutant versus IDH-wildtype gliomas. The architecture was an 18-layer residual design adapted to volumetric convolutions, trained from random initialization [36]. The two input channels were a robustly normalized T1c volume and the binary ET mask. Intensity normalization used the median and interquartile range of nonzero voxels within each T1c volume, followed by clipping to [− 6, 6]. The volume and mask were cropped or zero-padded to a 96 *×* 96 *×* 96 voxel cube centered at the ET centroid. No interpolation of MRI voxels was performed during cropping; the CNN input therefore retained local anatomy and the segmentation geometry. Random flips along the three volume axes and small Gaussian perturbations of the normalized intensity channel were applied only during training.

The benchmark used the 416 independent baseline patients with `MUSE` eligibility and known IDH status (43 mutant and 373 wildtype), preserving the five preassigned, patient-disjoint, stratified outer folds. Each training partition was separated once into an internal model-fitting subset and a stratified validation subset containing 15% of that partition. Optimization used AdamW (learning rate 10^−4^, weight decay 10^−4^), weighted binary cross-entropy with the training-set ratio of negative to positive cases, batch size 2, and up to 100 epochs. Validation ROC-AUC controlled early stopping after 15 epochs without improvement; the best validation checkpoint was evaluated once on each held-out outer fold. No pretrained weights or external training cohort were employed. The pooled out-of-fold predictions produced the reported classification metrics; sensitivity, specificity, and balanced accuracy used a fixed decision threshold of 0.5. The CNN’s sigmoid scores were not clinically calibrated probabilities.

The independent CNN analysis supplied an image-based prediction for each eligible patient. For direct descriptive correspondence, we also assessed its already computed out-of-fold scores on the subset of 375 subjects shared with the native `MUSE` and oriented GLCM study. This restriction changes the evaluation population but does not retrain the CNN or alter its original outer-fold predictions. Native `MUSE` curves and GLCM observations were inspected *after* classification in selected cases to contextualize spatial organization. The CNN and variographic field remained independent computational outputs; no radiomic or `MUSE` measurements entered the network architecture. Clinical interpretation distinguishes molecular IDH status from benign versus malignant diagnosis or WHO grade.

## 4. Experimental Results

The experimental evaluation examines how original `MUSE` direction-dependent semivariance measurements relate to multiscale radiomic statistics and to the molecular annotations of the same glioma cohort. The analysis first verifies the source archive and physical correspondence between representations, then evaluates between-patient agreement and voxel-pair support. The subsequent experiments investigate IDH associations across native direction and lag coordinates, illustrate representative spatial patterns, and report independent CNN classification with case-level post-prediction characterization.

### 4.1 Technical feasibility and complete-field integrity

The original `MUSE` archive contained 136,448 experimental directional curves and 1,151,687 original directional lag observations across 416 baseline cases. The subset of 375 patients with directional GLCMs contained 123,000 curves and 1,075,217 lag observations. Every native measurement could be traced to the frozen subject source JSON using source-hash verification. Four ET-positive acquisitions had technical `MUSE` failures, and examinations with no enhancing voxels could not yield an ET-restricted representation. No post hoc removal was applied solely because an observed variogram was unusually anisotropic. The individual 328-curve fields are the primary result of `MUSE`, and all tabulated scalar statistics in this section quantify aspects of their comparison without replacing the fields.

In the 416-subject baseline cohort, the available conventional analysis included 396 grade-4 tumors; 43 cases were IDH-mutant and 373 were IDH-wildtype. The paired 375-subject cohort comprised 33 IDH-mutant and 342 wildtype tumors. As the ROI was defined by enhancement, the analyzed cohort was strongly enriched for enhancing grade-4 gliomas. The ET-absent exclusion is therefore also a biological selection criterion, not merely an acquisition failure.

### 4.2 Geometric compatibility between native MUSE and GLCM

The 375-patient paired dataset yielded 29,250 requested GLCM offset observations (78 per case), of which 28,683 met the GLCM support gate. Under the primary angle–lag–`MUSE`-support thresholds, 9,371 offsets (32.7% of supported GLCM offsets) had at least one geometrically matching `MUSE` measurement; under the strict criteria, 2,254 (7.9%) were matched. All 78 GLCM orientations lay within about 2.8^°^ of some `MUSE` direction, indicating that physical-lag compatibility was a primary determinant of missing matches. Across the selected matches, only 4,581 unique native `MUSE` curves were involved, approximately 3.7% of all available paired-cohort curves. Unmatched curves were retained and cannot be equated with new or exclusive biological information.

Geometric compatibility diminished as the physical length of the tested offset increased. Of supported step-1 offsets, 4,116 of 4,864 (84.6%) met the primary match definition. The corresponding figures were 2,618/4,861 (53.9%) for step 2, 1,122/4,852 (23.1%) for step 3, 1,129/4,809 (23.5%) for step 5, 35/4,715 (0.74%) for step 8, and 351/4,582 (7.66%) for step 11. The non-monotonicity reflects the original discrete lag configurations and available support.

At the 9,371 accepted matches, the Pearson correlation between original `MUSE` semivariance and GLCM contrast divided by two was *r* = 0.954 (Spearman 0.962). The median within-patient Pearson correlation among patients with sufficient matches was 0.951, with a patient-resampled 95% interval of approximately [0.948, 0.954]. After removing patient and exact physical-offset-distance effects, the correlation across orientations was 0.846 (cluster-resampled interval [0.828, 0.863]). The median difference between half GLCM contrast and `MUSE` semivariance was approximately −0.017, with median absolute discrepancy 0.043; these numerical discrepancies reflect nonidentical estimators and are not measurement errors against an assumed ground truth.

### 4.3 Cohort-level agreement between complete native fields and oriented GLCMs

Among 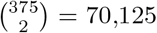 possible between-patient combinations, 31,599 (45.1%) satisfied the predetermined volumetric, GLCM, and native-point coverage gates. The rank correlation between the matched-original-`MUSE`-field RMSE and the full 468-feature GLCM distance was *ρ*_S_ = 0.862 (Fig. 5). The association between representations remained stable when examined across repeated patient-subset selections. Under repeated selection of 80% of patients, the descriptive correlation generally ranged from about 0.849 to 0.874; leave-one-patient-out values were approximately 0.859–0.865. Pairwise observations share patients, and these intervals describe patient-subset stability and have no interpretation as independent-pair confidence intervals.

**Figure 5:**
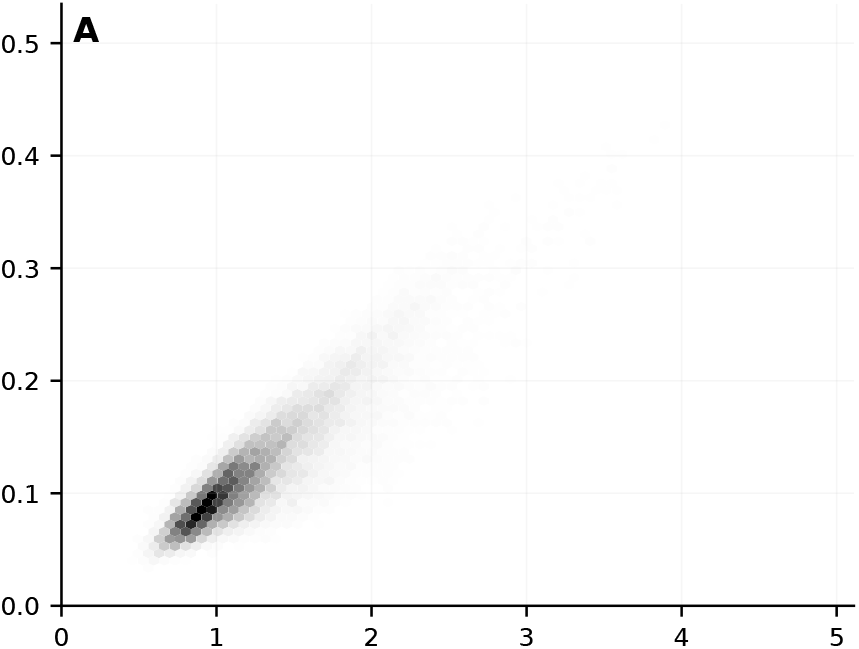
Between-tumor comparison over 31,599 eligible patient pairs. Standardized dissimilarity across 468 original directional GLCM measurements (horizontal axis) and root-mean-square difference between physically matched original `MUSE` semivariance samples (vertical axis). Gray density indicates the number of eligible comparisons in each hexagonal bin. Family-specific rank correlations are reported numerically in Table 2.

**Figure 6:**
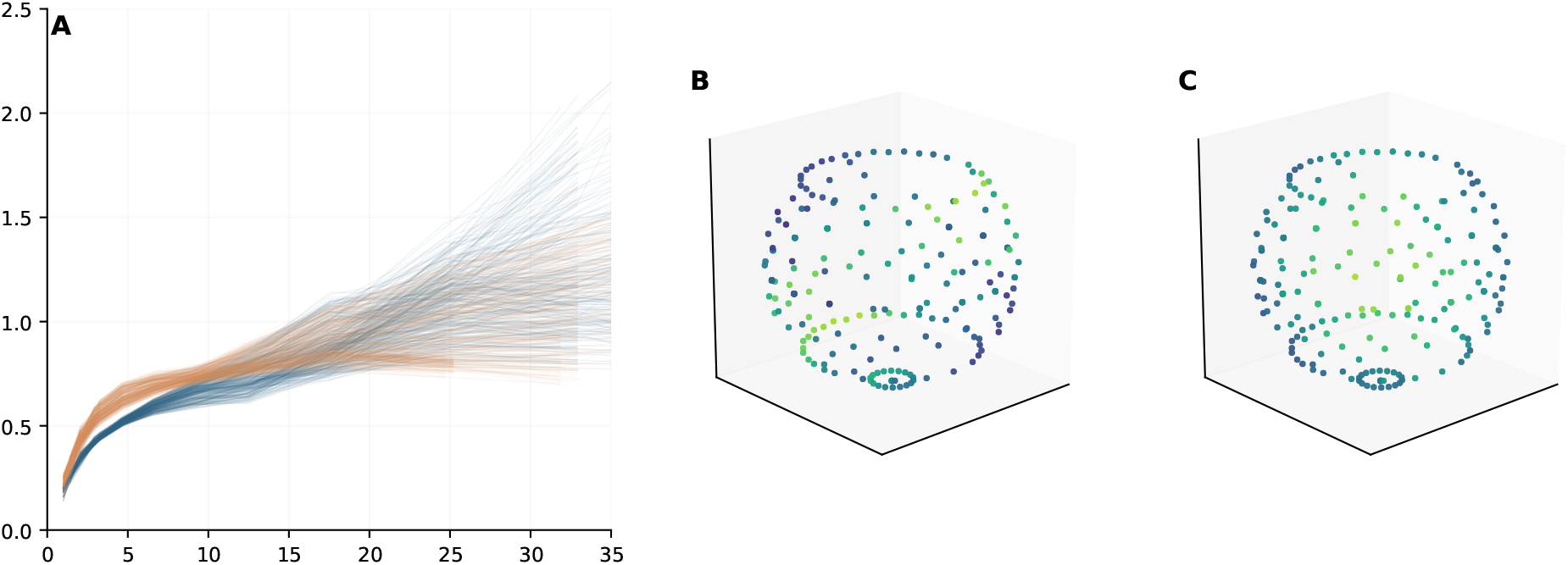
Original directional variograms and 3D sampled orientation geometry for two WHO grade-4 glioma cases. **(A)** Experimental semivariance (vertical axis) against native physical lag in millimetres (horizontal axis), with 328 original curves per patient for UCSF-PDGM-0531 (blue) and UCSF-PDGM-0096 (orange); only samples supported by at least 100 voxel pairs are displayed. **(B**,**C)** Original axial orientation vectors for the same patients at the exact native lag of 17.4744 mm. The 3D point coordinates are the measured `MUSE` direction components; color represents original semivariance on a shared scale (dark blue low, yellow high). The 3D panels show native sampled directions, without interpolation or ellipsoid fitting.

The relationship differed across the six GLCM feature families. Contrasts based on intensity differences were most concordant with the native variographic dissimilarity (GLCM contrast: *ρ*_S_ = 0.925; dissimilarity: 0.921). GLCM correlation and homogeneity yielded 0.831 and 0.804, whereas entropy and energy yielded 0.652 and 0.606 (Table 2). These results are consistent with a shared second-moment dependence and with the distinct distributional properties measured by co-occurrence entropy and energy. They do not demonstrate that the less concordant feature families contain independently useful clinical information.

**Table 2.** Spearman concordance between the complete-field MUSE intertumor dissimilarity diagnostic and GLCM dissimilarities computed from different feature families, across 31,599 eligible, nonindependent patient pairs.

| GLCM representation | $\rho_S$ with MUSE | Number of oriented measurements |
| --- | --- | --- |
| Contrast | 0.925 | 78 |
| Dissimilarity | 0.921 | 78 |
| Correlation | 0.831 | 78 |
| Homogeneity | 0.804 | 78 |
| Entropy | 0.652 | 78 |
| Energy | 0.606 | 78 |
| All six families | **0.862** | **468** |

### 4.4 Sampling support explains important apparent discordances

The original diagnostic selection found 15 `MUSE`-close/GLCM-far pairs under a 20th/80th-percentile comparison, three of which were exported as case examples. Three additional both-close and three both-far pairs were exported as controls; therefore, the nine selected examples are not nine discordances. The three original `MUSE`-close/GLCM-far examples were UCSF-PDGM-0126/0291, 0340/0430, and 0095/0430. Between 62.9% and 73.0% of their squared robustly scaled GLCM distance came from entropy and energy, and between 61.7% and 72.0% from the largest voxel step. These percentages describe the selected numerical distance, not explained biological variance.

The comparison 0340/0430 illustrates the problem: at one step-11 offset, 989 versus 121 contributing voxel pairs yielded entropy 8.529 versus 6.718 bits and energy 0.00344 versus 0.01185. The sparse distribution is especially sensitive to the finite number of distinct observed transitions; because the implementation symmetrized counts, the entropy upper bound for 121 original pairs is log_2_(242) ≈7.92 bits, not log_2_(121). Symmetric counting does not render the two sets of spatial pairs statistically independent.

When the previously extracted GLCM feature entries were masked unless both tumors met progressively stricter support gates, representation concordance rose from 0.862 at 100 pairs to 0.871, 0.875, 0.879, and 0.879 at the 200-, 300-, 500-, and 1,000-pair thresholds (Table 3). None of the three originally exported discordant pairs remained in the `MUSE`-close/GLCM-far category at a gate of 200 or more pairs. Nevertheless, five *other* discordant pairs remained at the 500-pair threshold, so the evidence concerns the instability of the original examples and does not establish universal equivalence. Excluding entropy and energy increased `MUSE` –GLCM concordance to 0.913; using only contrast and dissimilarity yielded 0.930. Both operations change the statistical construct and are reported as diagnostic sensitivity checks.

**Table 3.** Sensitivity of between-tumor rank concordance to the minimum GLCM voxel-pair support. The same 31,599 eligible patient pairs were used; low-support oriented entries were masked, and no image features or `MUSE` variograms were recomputed.

| Minimum offset support | MUSE –GLCM $\rho_S$ | Original three discordances retained |
| --- | --- | --- |
| 100 | 0.862 | 3/3 |
| 200 | 0.871 | 0/3 |
| 300 | 0.875 | 0/3 |
| 500 | 0.879 | 0/3 |
| 1,000 | 0.879 | 0/3 |

### 4.5 Molecular association localized to original 3D direction–lag measurements

The primary patient-level association analysis evaluated 2,508 supported *original* direction–lag cells within 364 grade-4 tumors. The global permutation test after conventional clinical and morphologic adjustment was significant (*p* = 0.00050). In this model, 894 cells retained an FWER-adjusted *p <* 0.05 after maxT correction over the tested spatial family. Most were at native physical lags approximately 1 mm to 5 mm; the signed IDH-mutant-minus-wildtype test statistics were negative, compatible with lower conditional semivariance for IDH-mutant tumors in those locations (Table 4). These test scores are standardized residual statistics and should not be interpreted as direct effect sizes.

**Table 4.** Association between IDH status and original MUSE semivariance at 2,508 exact 3D direction–lag cells. Models were fitted within the WHO grade-4 subgroup (*n* = 364; 24 IDH-mutant); 2,000 patient-wise stratified permutations calibrated the global statistic and maxT FWER.

| Covariate adjustment | Global permutation $p$ | Significant native cells | Minimum cell FWER $p$ |
| --- | --- | --- | --- |
| Conventional only | **0.00050** | **894** | 0.00050 |
| Conventional + GLCM PC5 (primary) | 0.2564 | **0** | 0.1064 |
| Conventional + GLCM PC10 (sensitivity) | 0.0720 | 1 | 0.0395 |

After the *primary* addition of five PCs of the full set of 468 oriented GLCM measurements, the global patient-permutation *p* increased to 0.256, and no cell survived the maxT adjustment. Under the sensitivity specification with ten oriented-GLCM components, the global *p* was 0.072, with one coordinate nominally passing within-model maxT correction (*p*_FWER_ = 0.0395): plane 13, local direction 1, original lag 17.4744 mm. The same coordinate had *p*_FWER_ = 0.1064 in the five-PC primary model. Consequently, that cell was *not* treated as a robust independent clinical finding. The three tests and their clinical interpretation are summarized in Table 4. The permutation procedure preserved whole-patient labels across the correlated direction–lag cells; 7 of 18 age–volume–sex strata contained both IDH groups, limiting the available conditional exchanges.

A prior secondary analysis of compact fitted `MUSE` summaries had found conditional associations of local spatial coherence with IDH and overall survival under conventional adjustment. In the 375-patient GLCM-matched cohort, nested five-fold IDH classification yielded ROC-AUC 0.880 using conventional variables, 0.915 after adding that short-range summary, and 0.937 using conventional plus multiscale radiomics/GLCM features. Adding the same `MUSE` summary to the radiomic model yielded 0.933; including three compact `MUSE` fitted summaries yielded 0.938. Under regularized Cox modeling, held-out C-index was 0.7010 for conventional plus radiomics/GLCM and 0.7011 after adding the short-range summary. These *secondary summary-level comparisons* do not test the full native 3D field and are not used as evidence that a compact `MUSE` representation exhausts its spatial interpretability.

### 4.6 Full-field examples of spatial-scale and direction-dependent behavior

Original experimental curves of the 29 tumors in the predefined case-study set were inspected together with their 468 oriented GLCM values. Three of the 15 candidate opposite-IDH pairs fell below the 20th percentile of GLCM distance, and none of those three were farther than the 80th percentile of global `MUSE` field distance. Within this selected set, the two distances were strongly associated (*ρ*_S_ = 0.907). The examples therefore illustrate how original curves can be read and compared; they do not constitute a selective demonstration of extra molecular discrimination.

The pair UCSF-PDGM-0089 (IDH-mutant) and UCSF-PDGM-0487 (IDH-wildtype) had a nearly identical ET volume (volume ratio 1.012). Its GLCM distance was 0.693, at the 2.8th percentile of the eligible paired-cohort distribution, while the native `MUSE` comparison RMSE was 0.076, at the 13.7th percentile.

Original matched directions nevertheless showed a scale-dependent relative ordering: the mean semivariance difference (mutant minus wildtype) was about −0.070 at 1–3 mm, +0.075 at 6–10 mm, and +0.097 at 10–15 mm. These values are descriptive summaries of matching *original* curve samples, not new derived patient biomarkers. The GLCM also changes at corresponding offsets; the particular benefit shown here is that the native continuous-lag curves make the change of sign and its physical scale directly legible.

The second pair, UCSF-PDGM-0531 (IDH-mutant) versus UCSF-PDGM-0096 (IDH-wildtype), showed semivariance differences of approximately −0.118 at 3–6 mm, −0.090 at 6–10 mm, −0.018 at 15–20 mm, and +0.128 at 20–30 mm. The latter range was supported by 300 directly paired native directions. In this *specific GLCM protocol*, the maximum sampled offset is 19.05 mm; the original `MUSE` curves therefore expose longer-range behavior not represented among the extracted GLCM offset measurements (Fig. 6). This statement concerns the chosen extraction schedule and is not a fundamental inability of GLCMs to sample larger spatial offsets.

At the exact lag of 17.474 mm, the 224 original directions available per tumor show visually different directional semivariance patterns. For 0531, the 10th–90th percentiles across those native directions were approximately 0.764–1.024, versus 0.820–1.008 for 0096. Figure 6 displays the original 3D axial direction vectors at the exact lag, with semivariance represented by point color. The underlying measurements are preserved without angular interpolation. The display is a practical example of the access that a native variographic field provides to the joint geometry of distance and direction. It does not imply a unique molecular signature, because every illustrative pair was constructed to have opposite IDH status and no robust cohort-level incremental association remained after primary GLCM adjustment.

### 4.7 Independent convolutional classification and post-prediction spatial characterization

The standalone 3D residual CNN was evaluated on 416 independent patients, including 43 IDH-mutant tumors. Pooling the test predictions from the five patient-disjoint outer folds yielded a receiver operating characteristic area under the curve (ROC-AUC) of 0.792 and a precision–recall area under the curve (PR-AUC) of 0.414. The proportion of mutant cases in this population was 10.3%. With a fixed sigmoid decision threshold of 0.5, balanced accuracy was 0.742, sensitivity 0.744, and specificity 0.740. Across the five test folds, individual ROC-AUC values ranged from 0.712 to 0.914; the spread is relevant to the limited number of mutant patients in each fold (8 or 9). The model used MRI intensities and an ET mask only; its results therefore constitute an independent imaging classification benchmark (Table 5).

**Table 5.** Standalone three-dimensional residual CNN classification of IDH mutation status. Metrics were computed from patient-disjoint out-of-fold sigmoid predictions. The common-cohort results restrict the same precomputed predictions to the 375 subjects also included in the directional `MUSE`–GLCM analysis; no model was refitted. Sensitivity, specificity, and balanced accuracy use the fixed threshold of 0.5.

| Evaluation quantity | Baseline cohort | MUSE –GLCM intersection |
| --- | --- | --- |
| Patients, $n$ | 416 | 375 |
| IDH-mutant, $n$ | 43 | 33 |
| IDH-wildtype, $n$ | 373 | 342 |
| ROC-AUC | **0.792** | 0.808 |
| PR-AUC | 0.414 | 0.396 |
| Balanced accuracy, threshold 0.5 | 0.742 | 0.757 |
| Sensitivity, threshold 0.5 | 0.744 | 0.758 |
| Specificity, threshold 0.5 | 0.740 | 0.757 |

The independent model can also be inspected on the tumors used for native spatial comparisons. The out-of-fold mutant-class scores for UCSF-PDGM-0531 and UCSF-PDGM-0096 were 0.881 and 0.030, respectively, consistent with their mutant and wildtype labels. Their native `MUSE` fields show distinct direction- and lag-dependent semivariance patterns, including the longer-range relative change described in Section 4.6. For UCSF-PDGM-0089, an IDH-mutant case, the CNN score was 0.096, giving a wildtype classification at the prespecified threshold; the matched wildtype case UCSF-PDGM-0487 received a score of 0.121. Their original `MUSE` curves remain directly interpretable across physical lag distances irrespective of whether the CNN label was correct. The spatial patterns are an independent description of the image domain and provide no evidence that a CNN classification error can be corrected from these illustrative cases. This pairing demonstrates the intended post-prediction use: a molecular classification accompanied by a traceable spatial characterization of the corresponding tumor.

## 5. Discussion

The findings are considered in terms of shared spatial information, direct interpretability of the original directional field, reliability of sparse-offset texture estimates, and the limits of molecular association. The implications for further biomedical validation follow from these observed results.

### 5.1 A framework of shared spatial information with different interpretive representations

This study demonstrates that 3D experimental variograms and multiscale oriented GLCMs capture overlapping aspects of MRI spatial organization. This finding is supported by two measurements based on separate comparison procedures. First, local semivariance and half contrast correlate strongly at physically comparable offsets. Second, original-field `MUSE` tumor dissimilarities and GLCM tumor dissimilarities show strong rank concordance across more than thirty thousand eligible between-patient comparisons. This agreement is an expected property of two statistical descriptions derived from intensity differences in a common ET domain. It should not be presented as a methodological limitation or as evidence that the native field can be discarded.

Each representation preserves different observational coordinates and statistical properties. `MUSE` retains a physical separation variable, an axial direction vector, the original semivariance estimate, and the support for every lag, allowing the investigator to inspect the shape and direction-dependent evolution of measured spatial dependence. The six GLCM measures summarize the joint distribution of quantized intensity pairs at a prescribed offset; contrast is closely related to the second moment, whereas entropy and energy depend on how co-occurrence mass is distributed. The wide range of measured intertumor concordance across GLCM families is consistent with this distinction. It does not license the assertion that a residual correlation gap is an estimate of independent biological information.

### 5.2 What the complete native field adds to biomedical interpretation

The plane-specific illustration in Figs. 3 and 4 clarifies the relationship between measurements and geometric summaries. Four experimental curves supply directly observed scale dependence within one plane, whereas fitted planar and 3D ellipses summarize preferred continuity extents in a compact geometric form. The ellipsoid is therefore an accessible companion visualization of anisotropy, and its fitted semi-axes should not be interpreted as independent measurements or as a substitute for the native 328-curve field. Because the supplied illustration lacks a verified UCSF case identifier, it provides methodological context and carries no patient-level molecular interpretation.

The original field supports questions that can be phrased and visualized in physical terms: at which separation do two tumor regions cease to resemble one another; do certain 3D orientations show greater semivariance than orthogonal orientations; and does the relative pattern change with scale? The original semivariance curves from the selected case pairs illustrate how these properties can be inspected in physical coordinates. In case 0089/0487, the ordering of the semivariance differences changes between short and intermediate separations even though both global GLCM and `MUSE` dissimilarities are low. In case 0531/0096, the relative ordering changes at long separations, some beyond the largest GLCM offset specified by the analysis protocol. At 17.474 mm, native 3D direction vectors expose different angular structures alongside the full experimental semivariance curves.

These observations document *interpretive access*, defined here as direct traceability from a spatial claim to its measurement coordinates and physical support. Clinical decision performance, pathological correspondence, and robustness to alternative GLCM offset schedules require additional investigation. In particular, direction-resolved GLCM measurements contain explicit orientations as well, and the conclusions depend on the specific set of 78 offsets extracted. A rigorous downstream reader should consider spatial patterns together with the original images and segmentation to evaluate possible effects of mask morphology and heterogeneous enhancement. The original T1c and ET volumes for patient UCSF-PDGM-0531 are shown in Figure 1; direct spatial registration of each variogram direction with the corresponding intratumoral structures remains a target for future validation.

### 5.3 Support sensitivity is essential to claims of complementarity

Finite voxel-pair support is not merely a technical nuisance. Shape and volume determine which separations occur within an enhancing mask and how many distinct voxel pairs can contribute to a texture estimate. At long offsets, a symmetrized GLCM estimated from few pairs can show inflated energy or constrained entropy even in the absence of a stable biological difference. The loss of the original three `MUSE`-close/GLCM-far exemplars under stricter saved support thresholds illustrates how a visually persuasive numerical discordance can arise from measurement design. Support-aware interpretation is therefore required for comparisons between the representations.

Similarly, only a subset of GLCM offsets could be paired exactly to original `MUSE` lags and angular directions within the selected geometric tolerances. A nonmatchable offset does not establish information missing from GLCM. `MUSE` and GLCM also aggregate voxel pairs differently. Comparisons at equal distance or approximate orientation are useful diagnostics, but equating their estimates without matching intensity representation and pair sets would be incorrect. These considerations are relevant to any prospective standard for joint radiomic–geostatistical analysis.

### 5.4 Independent prediction and descriptive interpretation

The CNN establishes a patient-level image-classification reference that can accompany the directional spatial profile measured by `MUSE`. Its voxel-based residual layers learn predictive representations from the T1c crop and ET geometry, while the archived `MUSE` field exposes a physical lag axis, 3D orientations, semivariance, and voxel-pair support. Reporting the two outputs for the same case makes both a molecular prediction and a directly inspectable account of spatial signal organization available for biomedical assessment. Such post-prediction characterization has a different evidential meaning from attribution techniques that estimate which voxels or learned representations caused a network output. No internal CNN attribution map, causal explanation, or clinical decision improvement is established by the present experiment.

The standalone CNN obtained a cohort-level ROC-AUC of 0.792, with substantial variation across the five external test folds of the internal validation protocol. The 375-case subset ROC-AUC of 0.808 is a descriptive restriction of predictions produced by models trained on the larger 416-case cohort. The CNN and radiomic classification experiments differed in preprocessing, feature domain, fitting protocol, and, in some cases, evaluation population. Their AUC values are therefore not a paired head-to-head estimate of model superiority. The complete `MUSE` field was evaluated principally through physical interpretation and support-aware comparison with GLCMs, not as a predictive replacement for the CNN. A prospective clinical evaluation could test whether access to the spatial description changes readers’ assessment, error detection, or downstream decisions beyond the image and classifier score alone.

### 5.5 Clinical associations and the distinction between association and utility

The native-field grade-4 IDH analysis detected a broad association with semivariance under conventional adjustment. The predominantly short-range and negative signed cell scores are compatible with a shift in short-range spatial coherence, a biologically interpretable aspect of intratumoral MRI organization. The strong attenuation after adjusting for directional GLCM components is equally informative. No native direction–lag cell remained significant under the primary five-PC oriented-GLCM model, and the global IDH test was not significant. A single significant cell under the ten-PC sensitivity model depended on the adjustment specification. The study therefore does not substantiate an independent `MUSE`-derived IDH marker beyond the radiomic representation tested.

Prior summary-level analyses also associated a `MUSE` short-range measure with IDH and overall survival after conventional adjustment. The same measure was strongly reconstructable from harmonized multiscale radiomics in held-out folds and did not improve internally validated IDH AUC or survival C-index once the enriched radiomic model was available. These secondary analyses characterize the overlap of specific scalar summaries; they neither replace the 328 original `MUSE` curves nor define the clinical performance of the complete field. Molecular subtype is a clinically relevant annotation, but it is not synonymous with the directly observable geometric structure of enhancement, and the 2021 WHO framework integrates multiple molecular and histologic variables [1].

The opposite-IDH case pairs provide illustrative comparisons; molecular association is evaluated at the patient-cohort level. All selected tumors were grade 4; they cannot be interpreted as examples of differing WHO severity grade. Survival comparisons require censoring-aware adjusted analyses across patients. A demonstrable clinical *benefit* would require an independently validated increase in prognosis, prediction, stratification, or clinical decision quality, ideally in multicenter data and with explicit controls for treatment and selection. The present results instead justify a more precise claim of descriptive and physical interpretability with a measured, and partly shared, relationship to molecular annotation.

## 6. Conclusions, Limitations and Future Work

We introduced and evaluated an integrated radiomic–geostatistical workflow that preserves the original 328-curve 3D `MUSE` experimental representation per enhancing glioma and places it in explicit correspondence with multiscale directional GLCM statistics. The paired experiments demonstrate strong shared second-order spatial information, family-dependent differences in co-occurrence texture, and substantial sensitivity of apparent discordance to spatial pair support. Original patient examples show the value of retaining scale and 3D orientation as directly inspectable physical coordinates, including behavior beyond the specific GLCM offsets extracted. In the WHO grade-4 subgroup, a widespread `MUSE` –IDH association under conventional adjustment was largely accounted for by oriented GLCM components, and no robust incremental clinical association was established in the primary adjusted analysis. An independent 3D CNN provided out-of-fold IDH classification (ROC-AUC 0.792 over 416 patients), and its predictions were inspected alongside the native spatial measurements in selected tumors. This joint reporting provides a classification outcome accompanied by physically interpretable tumor heterogeneity measurements, while clinical benefit from such post-prediction characterization remains to be established. The framework thus offers a traceable basis for complementary image classification and spatial tumor description.

### Limitations and future work

Several limitations define the external validity of the study. The source is one public imaging collection, with no independent institutional validation and no test–retest acquisition. The ET-restricted analysis excludes 75 examinations without enhancement and consequently represents predominantly grade-4 tumors; the native-field IDH analysis includes only 24 mutant cases, limiting power and exchangeability within the stratified permutations. Small or elongated tumor regions provide few voxel pairs at large offsets, which affects both `MUSE` and GLCM estimates. Normal-score and GLCM discretization choices, directional bandwidth, angle tolerances, and selected offset lengths affect the quantitative comparison. The original `MUSE` field is fully available, but the global between-tumor RMSE weights lag–direction observations according to coverage and cannot summarize every scale and orientation uniformly. The preselected clinical examples were examined post hoc, and the full spatial association test was developed in the course of the study; it is exploratory with no independent preregistered confirmation.

Adjustment by five or ten principal components of oriented GLCM features is deliberately finite-dimensional and cannot guarantee conditional independence from all 468 original oriented GLCM measurements, let alone arbitrary nonlinear transformations. The native 3D direction–lag coordinates are strongly dependent, and the number of candidate cells is not the number of independent biological signals. Permutations within age–volume–sex strata rely on conditional exchangeability; only seven strata contained both IDH classes. Tumor mask perturbation, repeated-image assessment, and scanner-to-scanner reproducibility remain untested in this analysis. A representative anatomical and ET overlay was generated from the original NIfTI image for UCSF-PDGM-0531; corresponding anatomy-to-direction inspection was not systematically performed for every selected case. The CNN was trained from scratch on a small and imbalanced single-cohort dataset with 8–9 mutant cases per outer test fold; we report internal out-of-fold discrimination without independent-center evaluation or score calibration. The ET mask was supplied as a CNN input, so its classification cannot be attributed to intensity patterns alone. The classifier and radiomic models were not matched in every training and preprocessing detail; comparing their reported AUC values does not establish predictive superiority. Finally, there is no prospective or external demonstration that the combined reporting of CNN predictions and `MUSE` spatial fields improves clinical decisions or patient outcomes.

These limitations motivate three connected next steps: evaluate stability of the original directional field under controlled mask perturbations and acquisition variation; extend the analysis to multiple tumor compartments and modalities where comparable spatial support exists; and test predefined, interpretable native-field hypotheses in independently annotated clinical cohorts. Future work can assess CNN and `MUSE` observations together in blinded clinical reader studies, preserving the original physical variogram observations and evaluating their effect on interpretation separately from the predictive accuracy of the network. These extensions require new evidence and define subsequent validation objectives.

## Data Availability

All data produced in the present work are retained in the authors research archives and have not yet been deposited in a public repository

## Data availability

The publicly accessible UCSF-PDGM imaging collection, segmentation masks, and associated annotations are available through The Cancer Imaging Archive (dataset DOI: 10.7937/tcia.bdgf-8v37). All data produced in this work are retained in the authors’ research archives and have not yet been deposited in a public repository.

## Code availability

The open-source `MUSE` methodological implementation is publicly available at https://github.com/mariannamiola/muse. The MRI-specific scripts for data preparation, directional GLCM extraction, statistical evaluation, and standalone CNN benchmarking have not yet been deposited in that repository. The publicly accessible `MUSE` code and the study-specific analysis scripts are distinct components of the workflow.

## Ethics and consent

This is a retrospective computational analysis of an openly shared, de-identified public imaging dataset. Dataset governance and conditions of reuse follow the UCSF-PDGM/TCIA release [18]. No new human participant recruitment or intervention was performed.

## Use of generative artificial intelligence

During the preparation of this work, the authors used ChatGPT 5.6 (OpenAI) to assist with the revision of the work.

## A. Supplementary analysis and reporting details

### A.1 Native MUSE archive and GLCM settings

The source run used automatic plane generation and four sampled directions on each of 82 planes (328 3D curves per patient). The original export retained 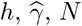 and source SHA256 for each point. The 375-case GLCM file was generated with steps {1, 2, 3, 5, 8, 11}, 13 3D axial offsets, normal-score rank mapping and a quantization bin width of 0.2. Six measures were available for every supported offset: contrast, dissimilarity, homogeneity, correlation, entropy, and energy. Invariant summaries were calculated only within physically equivalent offset-length classes. Source parameters were frozen before the comparison studies.

### A.2 Secondary radiomic overlap experiments

Earlier internally cross-validated models examined how much of selected `MUSE`-derived summaries could be statistically reconstructed from standard and multiscale radiomic features. The original omnidirectional curve comparison showed a held-out *R*^2^ of approximately 0.776 for its specified regression experiment; the published-in-project direction-dispersion target had *R*^2^ = 0.546 with conventional multiscale radiomics and up to 0.689 after additional directional GLCM information. These results assess *predictability of auxiliary `MUSE` targets*; they do not reconstruct or replace original `MUSE` measurements in the present study, and 1 − *R*^2^ is a regression diagnostic and has no direct interpretation as a proportion of exclusive biological information.

For completeness, the clinical benchmark based on one short-range `MUSE` summary obtained held-out ROC-AUC 0.880 using conventional clinical/morphologic variables and 0.915 after adding that summary. The radiomics/GLCM-enriched model achieved 0.937, versus 0.933 with the summary added and 0.938 after adding three compact fitted summaries. A ridge-Cox internal analysis of the paired cohort yielded C-index 0.7010 for clinical/radiomic variables and 0.7011 after adding the summary. This appendix records the secondary comparison and explicitly separates it from inference based on the original full directional field.

### A.3 Interpretation safeguards

The original `MUSE` archive, the GLCM table, and the clinical labels are different observational layers. The first describes spatial dependence conditional on an ET mask, the second a quantized transition distribution, and the third molecular, clinical, or time-to-event annotations. An association between any two layers is not evidence of anatomical causality, histological validation, or downstream clinical utility. The patient-level nature of the sampling unit, the influence of finite spatial support, and the reported null results after radiomic adjustment apply equally to all illustrated examples.

